# Investigating the Effect of Climate and Air Pollution on Prescription Uptake in the England

**DOI:** 10.64898/2026.02.13.26346258

**Authors:** John Tolladay, Christopher Yau

## Abstract

**Background:** Climate change is increasingly recognised as a threat to population health and healthcare systems, yet the effects of environmental variability on pharmaceutical prescribing remain poorly characterised in the UK. Using a wide array of open-source datasets, we examine the effect of environmental, geographic and socioeconomic factors on prescribing habits in England.

**Methods:** We linked monthly, practice-level prescribing data for England (2010–2025) to meteorological, air-quality, flooding and demographic datasets using spatial nearest-neighbour matching. Prescribing volumes for cardiovascular, respiratory and antibiotic medications were analysed using log-transformed outcomes in mixed-effects models with practice-level random effects, adjusting for region, seasonality, deprivation and temporal trends, using both continuous environmental measures and extreme-condition indicators. A complementary Bayesian hierarchical model jointly estimated the conditional effects of multiple correlated environmental exposures, with partial pooling across practices and support for distributed lag effects.

**Results:** In mixed-effects analyses, temperature showed the most consistent associations with prescribing, with higher temperatures linked to increased respiratory and cardiovascular prescriptions and reduced antibiotic use, while rainfall, flooding and most pollutants had small or negligible effects. Environmental predictors exhibited strong correlations, motivating multivariate modelling. Bayesian multivariate models confirmed temperature as the dominant environmental driver after adjustment for correlated exposures, with substantially larger variation attributable to regional and socioeconomic factors than to environmental conditions.

**Conclusions:** Temperature is the most consistent environmental determinant of GP prescribing in England, with higher temperatures associated with increased cardiovascular and respiratory prescribing and reduced antibiotic use. Rainfall, flooding and most air pollutants show little evidence of meaningful effects once seasonal and meteorological structure is accounted for. Environmental associations are modest in magnitude relative to persistent socioeconomic and regional drivers of prescribing, indicating that climate-related influences operate within broader structural determinants of healthcare utilisation. These results suggest that, at monthly timescales, prescribing demand is relatively stable to environmental variability, supporting a focus on long-term adaptation and surveillance rather than short-term demand shocks in climate-resilient healthcare planning.

## 1 Background

In recent years it has become clear that climate change poses threats to human health and the provision of medication. Preliminary studies and literature reviews have shown that more research is required to understand the impacts of climate change on healthcare systems [1, 2]. A recent Health and Climate Adaptation Report by National Health Service (NHS) England states that it is critical to empower health professionals with the tools and knowledge to integrate climate data into decision-making to build resilience in the healthcare system [3]. Several studies have been done into the effects of climate change on public health, with most finding that a general increase in cardiovascular, respiratory, infectious and mental-health related conditions will increase based on climate projections [4, 5, 6, 7]. In particular, recent work in the UK highlights that temperature extremes can drive increases in general practitioner (GP) consultations, hospital admissions and prescriptions, with disproportionate effects among older adults [8]. While some attention has been given to environmental effects on the supply and transport of medications [9, 10], less attention has been given to the effect on prescription volumes.

A study of datasets from Switzerland found that doctor visits and prescriptions are likely to increase with the changing climate, though statistical uncertainty limited the conclusiveness of their results [11]. Using United States population and climate data, the RAND Corporation modelled how climate change will influence future drug demand for four major chronic conditions. Their analysis projects increased demand for medications used to treat asthma, end-stage renal disease and Alzheimer’s disease, with a slight decrease for cardiovascular therapies, highlighting the importance of strengthening climate-resilient pharmaceutical supply chains [12]. Despite growing interest in climate–health interactions, the effects of climate factors on pharmaceutical prescribing in the United Kingdom (UK) have received little systematic study, underscoring the need for further investigation.

In this article we utilise freely available climate, air quality and prescription datasets to assess how prescriptions are affected by variability in environmental conditions over time. The aim is to determine whether there are relationships between factors such as temperature, rainfall, flooding and multiple forms of air pollution, and the uptake of prescriptions at the medical practice level. To do this, we combine meteorological observations from the UK Met Office for temperature and rainfall [13]; further rainfall data from the Department for Environment Food & Rural Affairs Hydrology Application Programming Interface (API) [14]; dated flood outlines from the UK Environment Agency [15]; surface level air pollution data from the UK Air Quality Reanalysis [16]; regional outlines from the Office for National Statistics [17]; and Index of Multiple Deprivation data from various government departments [18]. These datasets are linked to monthly practice-level prescribing data for England from the Open Prescribing [19] and NHS Digital [20] services, allowing us to quantify how environmental fluctuations relate to prescribing patterns across conditions and regions. This integrated dataset provides a foundation for evaluating potential climate–health–prescription relationships, with a focus on cardiovascular, respiratory and antibiotic treatments.

## 2 Method

### 2.1 Data collection

#### Prescription data

Monthly prescription data for England covering September 2020 to August 2025 were obtained from the Open Prescribing Application Programming Interface (API) at the individual practice level. Historical prescription data from August 2010 to December 2019 were downloaded in bulk from the NHS Digital prescribing service and subsequently aggregated by month and medical practice. Four analytical datasets were then constructed by filtering prescriptions using British National Formulary (BNF) codes representing different therapeutic groups: BNF code 02 (cardiovascular medications), BNF code 03 (respiratory medications), BNF code 0501 (antibiotics) and a combined dataset containing all prescriptions from these three categories. In total, data from 15,166 practices were included in the base dataset. Each practice was assigned to its corresponding geographic region using data from the Office for National Statistics, enabling regional-level analyses and adjustment for regional differences in prescribing patterns. English regions used in the study were: London, South East, South West, East of England, West Midlands, East Midlands, Yorkshire and The Humber, North West, and North East.

#### Weather data

Historical temperature and rainfall data were obtained from 37 Met Office weather stations across the UK. Monthly minimum and maximum temperature (^°^C), and total rainfall (mm) were extracted for all periods covered by each station. Additional daily rainfall measurements from a wider network of locations were retrieved via the Department for Environment, Food & Rural Affairs (DEFRA) Hydrology API and aggregated to monthly totals to ensure temporal alignment with the prescription and Met Office records. The Met Office stations provided data spanning varying time ranges, collectively covering January 1900 to September 2025, while the hydrology dataset extended from January 1957 to October 2025. Together, these sources fully encompassed the period for which prescription data was available. For each medical practice, the nearest Met Office and hydrology stations were identified using k-d tree spatial matching based on latitude and longitude.

#### Flood data

Flood exposure was characterised using a comprehensive dataset of historical flood outlines provided by the UK Environment Agency, containing mapped flood extents from major events dating back to the English Channel storm of 1703 through to flooding events in January 2025. Each flood polygon within the time period covered by the prescriptions dataset was simplified to reduce geometric complexity and buffered by 5 km to capture practices in close proximity to affected areas. Monthly flood indicators were then generated by assigning each polygon to all months spanned by its recorded start and end dates. Using British National Grid coordinates for all practice locations, spatial intersection tests were performed to determine whether a practice lay within the buffered flood extent for each month. This process produced a binary monthly flooding “flag” variable for each individual practice.

#### Pollutant data

Monthly particulate matter (PM) concentrations were obtained from the UK Air Quality Reanalysis (AQREAN) dataset, which provides gridded estimates of air pollutant levels across the UK. Separate files were collected containing hourly estimates of mass concentrations (*µ*g m^−^3) and daily air quality index (DAQI) values for the pollutants carbon monoxide (CO), nitrogen oxides (NO, NO_2_, NO_*x*_), ozone (O_3_), sulphur dioxide (SO_2_) and particulate matter (PM_2.5_, PM_10_). For each pollutant, the monthly maxima at the nearest grid point to each practice were calculated over the time period of the prescription dataset. Although DAQI values are integers, they were treated as continuous variables to reflect their role as a scale rather than discrete categories.

#### Index of multiple deprivation

Index of Multiple Deprivation (IMD) data were compiled from the 2015, 2019 and 2025 releases. For each year, unique local authority district (LAD) codes and names were extracted to form a unified set of districts appearing in any dataset. LAD centroids were obtained from files corresponding to the 2019 and 2024 boundaries. The 2015 IMD data had already been refactored to the 2019 boundaries. The only consistently available measure across all three IMD datasets was “Rank of Average Ranks”, where lower values represent increased deprivation. Ranks were converted to percentile scores within each year to allow comparability despite changes in LAD numbers and boundaries. These percentiles were then aligned with the monthly prescribing dataset. For each practice, the nearest LAD centroid was identified separately for each IMD release year using a k-d tree nearest-neighbour search. Each monthly prescription record was then matched to the IMD release closest in time. The practice-specific nearest LAD for the selected IMD year was then used to assign deprivation percentiles to all months.

#### Conditional flagging

Binary and categorical “flags” were generated to identify months with anomalous or extreme weather or pollutant conditions. For each weather variable, observations from each station were standardised using median absolute deviation (MAD) scaling. Standardised monthly values were assigned to “high”, “low”, or “median” categories, where “high” and “low” corresponded to z-scores greater than +1.0 or less than −1.0, respectively, with all remaining months classified as “median”. For each pollutant mass concentration, the monthly maximum value for each practice was similarly standardised using MAD scaling. Months with pollutant z-scores exceeding 1.5 were flagged as “high”. Where daily air quality index (DAQI) values were available, these were categorised directly using established thresholds: “low” (1–3), “moderate” (4–6), “high” (7–9), and “very high” (10).

### 2.2 Data pre-processing

All analyses were conducted using harmonised prescription-environment datasets constructed from monthly GP prescribing data linked to meteorological, hydrological, air-quality, flooding and demographic records. For each prescription code (BNF 02, BNF 03, BNF 0501 and a combined dataset including all three), the dataset was loaded and processed using a consistent workflow. Practices were retained only if at least 20 monthly observations were available for both prescription counts and relevant predictors. All continuous predictors (including DAQI values) were standardised by subtracting the mean and dividing by the standard deviation across all practices and dates.

Prescription item counts were log-transformed to reduce skewness and approximate a normal distribution, enabling effect estimates to be expressed as percentage changes. Log-transformation was chosen over standardisation to retain meaningful between-practice differences while ensuring interpretable results. Practices with very low prescription volumes (mean monthly items ≤10) were excluded. For each remaining practice, values below 10% of the practice-specific mean were removed to avoid spikes or sudden changes in prescriptions that were unrelated to environmental variables. The dates were converted to an integer count of months since the first observation in the dataset and standardised to provide a numerically stable time index, improving parameter identifiability and convergence in both the Bayesian and mixed-effects models.

The resulting datasets were then passed to individual mixed-effects models and to a Bayesian multivariate model. Pairwise correlations were also calculated between continuous predictors and DAQI values for all captured dates and practices, with a mean value across all practices used for analysis.

### 2.3 Mixed-effects models

Two sets of mixed-effects models were fitted to quantify associations between each environmental variable and monthly prescribing. The models were implemented in Python using the `mixedlm` function from the `statsmodels` package. Each predictor was analysed in a separate model to avoid confounding from correlations among environmental variables. All models used the log-transformed prescription count as the outcome, so that estimated coefficients can be interpreted as approximate percentage changes in prescribing volume.

Covariates included in all models were region and Index of Multiple Deprivation (IMD) percentiles. IMD was treated as a time-invariant, practice-level covariate by taking the mean value for each practice across all months. Region and month were included as fixed effects using sum-to-zero coding, with coefficients representing deviations from the overall mean. All models included practice-level random effects, with random intercepts and random slopes on the standardised time index to capture between-practice differences and practice-specific prescribing trends.

Outputs from all mixed-effects models were stored for comparison across environmental variables and prescription groups, including coefficient estimates and 95% confidence intervals, for both fixed- and random-effects contributions.

#### Flag-based models

Flag-based models used binary or categorical indicators derived from the environmental datasets, including high/median/low temperature and rainfall flags, high pollutant concentration flags, DAQI category group flags, and a binary flood indicator. Each model produced an estimated effect size and confidence interval for the relevant flag. These models compared “high” and “low” to “median” weather conditions and “high” to not “high” pollutant mass concentrations. DAQI values were compared separately for “very high” or “high” versus “moderate” or “low”, and for “very high”, “high” or “moderate” versus “low” categories.

#### Continuous-value models

Another set of models were fitted using the standardised continuous values for each environmental predictor rather than categorical flags. Each predictor was modelled separately using the same mixed-effects structure described above. Flood exposure was not included in these models because it was available only as a binary flag.

### 2.4 Bayesian model

A Bayesian hierarchical regression model was used to estimate the combined and conditional effects of multiple environmental variables on monthly prescribing. The model was implemented in Python using the `PyMC`, `numpyro` and `jax` packages. Unlike separate mixed-effects models, this approach simultaneously incorporates many continuous predictors and the binary flood indicator, accounting for correlations among predictors. For each prescription code, the dataset included globally standardised meteorological, hydrological, demographic and pollutant variables, and the raw binary flood indicator. Following removal of missing data, each prescription code dataset retained between 6,775 and 7,220 practices with more than 20 months of available data. To improve model stability, Index of Multiple Deprivation (IMD) percentiles were averaged over time to generate time-invariant, practice-level fixed effects. IMD values were additionally mean-centred within regions to mitigate confounding between regional effects and deprivation. To mitigate collinearity, strongly redundant or derived predictors (e.g., Daily Air Quality Index components) were excluded, while correlated variables capturing potentially distinct healthcare effects (e.g., minimum vs. maximum temperature, PM_2.5_ vs. PM_10_) were retained.

#### Detailed model specification

Let *i* = 1, …, *N* index general practices and *t* = 1, …, *T*_*i*_ index monthly observations within each practice. The response variable was the monthly number of prescriptions, denoted items_*it*_. To stabilise variance and allow multiplicative interpretation of effects, the outcome was log-transformed as

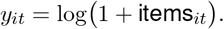

Let *r*(*i*) ∈ {1, …, *R*} denote the region associated with practice *i*, and let *m*(*t*) ∈ {1, …, 12} denote the calendar month corresponding to observation *t*.

Time was represented by a standardised numeric index *s*_*it*_, defined as the number of months since the first observation in the dataset and rescaled to have zero mean and unit variance.

The outcome was modelled using a hierarchical Bayesian regression of the form

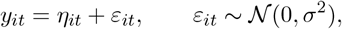

where the linear predictor is given by

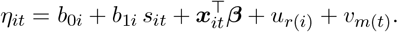

Practice-level effects were parameterised as deviations around population-level (global) effects. Specifically, a global intercept *α* and a global time slope *γ*_*t*_ were estimated for the population, and each practice was allowed to deviate from these via

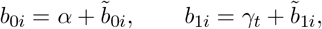

where the practice-specific offsets were modelled hierarchically as

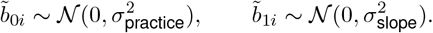

Random slopes on time were included only for practices with more than 50 observations. Both random intercepts and slopes were implemented using non-centred parameterisations with zero-mean constraints for identifiability. This hierarchical structure induces partial pooling across practices, stabilising estimates for practices with smaller sample sizes while allowing practices with more data to retain influence from their own observations.

The vector ***x***_*it*_ contained standardised continuous fixed-effect predictors, including environmental covariates and deprivation measures, with corresponding regression coefficients ***β***. Region effects *u*_*r*_ were modelled as partially pooled effects,

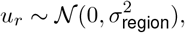

and seasonal effects *v*_*m*_ capture month-of-year variation and were modelled analogously as

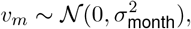

with both sets of effects constrained to have zero mean.

#### Prior specification

The global intercept *α* was assigned a weakly informative normal prior centred on the empirical mean of the log-transformed outcome. The global time slope *γ*_*t*_ was assigned a zero-mean normal prior with small variance, reflecting the expectation of gradual temporal change. Regression coefficients for standardised continuous predictors were assigned zero-mean normal priors, with scale parameters chosen to provide weak regularisation. Binary predictors were assigned broader zero-mean normal priors.

All standard deviation parameters governing hierarchical effects and residual variation (*σ, σ*_practice_, *σ*_slope_, *σ*_region_, and *σ*_month_) were assigned half-normal priors, providing regularisation while allowing substantial between-group variation when supported by the data.

#### Lagged predictors

To capture delayed effects of environmental exposures, predictors could optionally enter the model through distributed lag structures. When this option was used, lagged values (of maximum lag *L*) were first constructed within each practice. Rather than estimating a separate coefficient for each lag, the vector of lagged values was projected onto a low-dimensional polynomial basis prior to model fitting.

Specifically, for each predictor, the (*L* + 1) lagged values at time *t* are transformed into (*K* + 1) smooth basis variables using an orthogonalised Almon polynomial basis. These basis-transformed variables are then included directly as standard fixed-effect covariates in the linear predictor. This approach enforces smoothness across lags, reduces the effective dimensionality of the model and improves numerical stability. Importantly, because the model is parameterised in terms of basis coefficients rather than explicit lag weights, no explicit estimation of individual lag weights is required, and it is not necessary to impose identification constraints on their sum.

#### Inference

Posterior inference was performed using Hamiltonian Monte Carlo with the No-U-Turn Sampler. Eight chains were run, each with 3,000 warm-up (tuning) iterations followed by 2,000 posterior draws. Convergence was assessed using standard diagnostics, including trace plots and effective sample sizes. Posterior summaries were reported as means and 95% equal-tailed credible intervals, which provide stable, quantile-based summaries that are robust to skewness.

#### Interpretation

Model coefficients represent additive effects on the log-transformed outcome and therefore correspond to multiplicative effects on the original prescription count scale. For interpretability, these effects and their 95% credible intervals were converted to percentage increases or decreases.

## 3 Results

We present analyses in three stages: first describing correlations among environmental predictors, then estimating marginal associations using individual mixed-effects models, and finally quantifying conditional effects using a Bayesian multivariate framework.

### 3.1 Strong seasonal and physical dependencies among environmental exposures

Before fitting regression models, we examined the correlation structure among environmental predictors to assess collinearity and guide model specification. Many meteorological and air-pollution variables exhibit strong seasonal and physical dependencies, which can bias effect estimates if not properly accounted for. Understanding these relationships is therefore essential for interpreting results from both univariate and multivariate models.

For the 19 continuous predictors compared, 38 out of 171 possible variable pairs had absolute mean correlations greater than 0.5 across practices (Figure 1). Rainfall variables from separate data sources were reassuringly highly correlated with each other (*r* ≈ 0.98). Maximum and minimum temperatures were highly correlated (*r* ≈ 0.96), while PM_2.5_ and PM_10_ concentrations also showed strong positive correlation (*r* ≈ 0.92). Mass concentrations also showed expected high correlation with their Daily Air Quality Index (DAQI) counterparts. Remaining correlations were all below 0.69. Temperature variables were moderately positively correlated with ozone levels and negatively correlated with nitrogen-based pollutants. Rainfall variables showed weak associations with many air pollutants. Nitrogen-based pollutants were strongly intercorrelated and also correlated with carbon monoxide and sulphur dioxide, while ozone tended to have negative associations.

**Figure 1.**
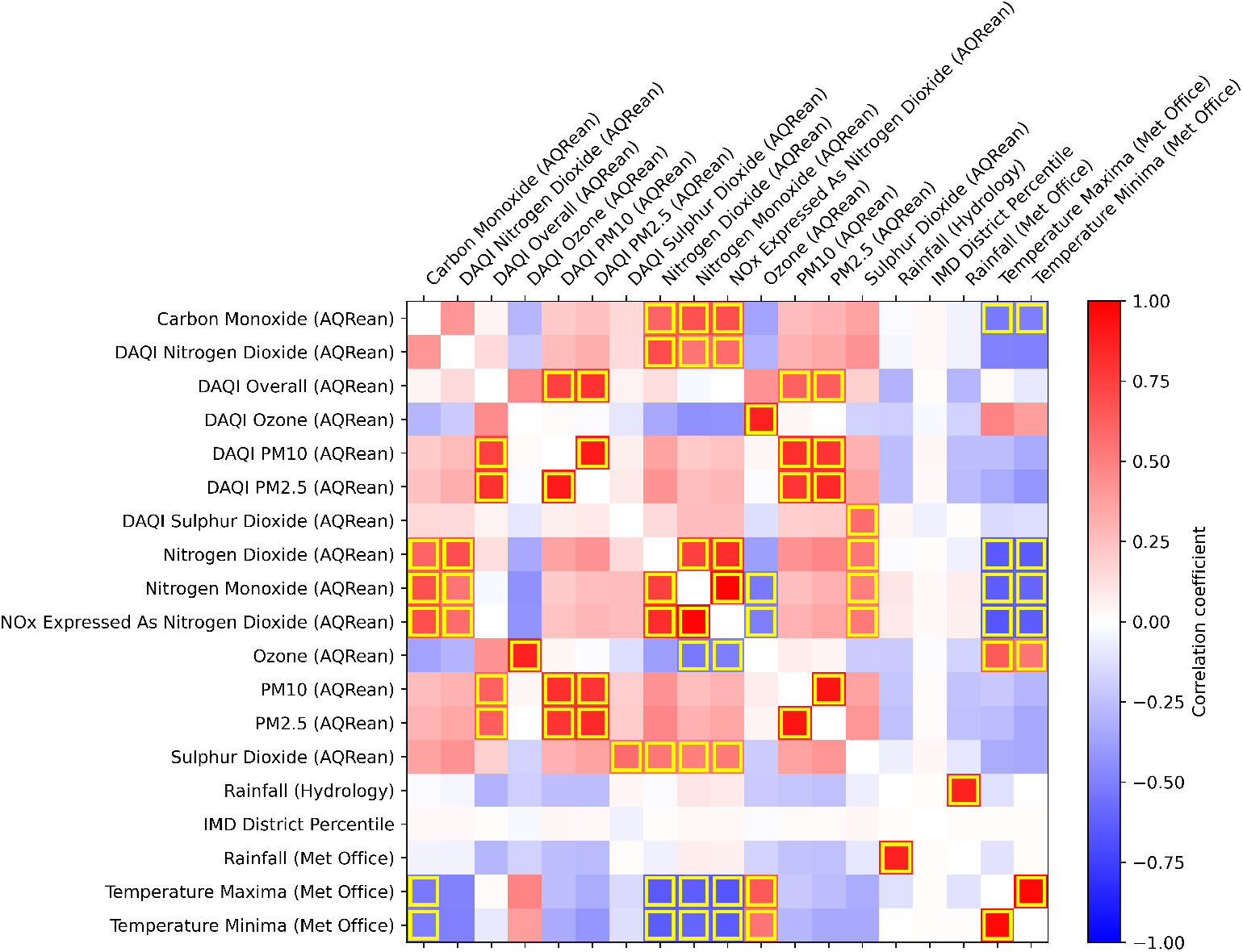
Pair-wise correlations between environmental predictors. Positive correlations are in red and negative correlations in blue, with absolute values > 0.5 marked with yellow boxes.

These correlations highlight the need for modelling approaches that can accommodate strongly related environmental exposures, motivating the use of both single-predictor mixed-effects models and a fully adjusted Bayesian multivariate framework with a reduced set of predictors in subsequent analyses.

### 3.2 Marginal environmental signals in prescribing behaviour

We first examined the associations between individual environmental predictors and prescription volumes using separate mixed-effects models. These models estimate the marginal relationships between each exposure and prescribing while controlling for region, deprivation, seasonality, and long-term trends. Although correlated predictors cannot be interpreted causally in isolation, these analyses provide an interpretable summary of how prescribing responds to variability in specific environmental factors.

#### Prescribing responses to anomalous and extreme conditions

We first assessed how prescribing behaviour changed during months characterised by unusually extreme environmental conditions. Such periods may capture non-linear responses, behaviour disruption, or system-level effects that are not well described by average changes in exposure. Flag-based models allow us to test whether prescriptions respond differently during anomalous weather events, high pollution episodes, or flooding compared to typical conditions.

Figure 2 summarises the coefficients for each flagged variable, expressed as average percentage changes in monthly prescription items. Months with unusually high minimum or maximum temperatures were associated with increases in prescriptions across all medication categories, ranging from 1.2–3.1%. Low-temperature months showed minimal associations for most categories (< 1% average change), although months with unusually low maximum temperatures were associated with 1.1–1.6% reductions in respiratory medication prescriptions. Associations with anomalous rainfall were generally weak, leading to < 1% changes in most prescription types, except antibiotics, which showed reductions of 0.8–1.4% during low rainfall months. Similarly, flooding had little effect on most prescription categories, but was associated with reductions of 2.7–3.7% in antibiotic prescriptions.

**Figure 2.**
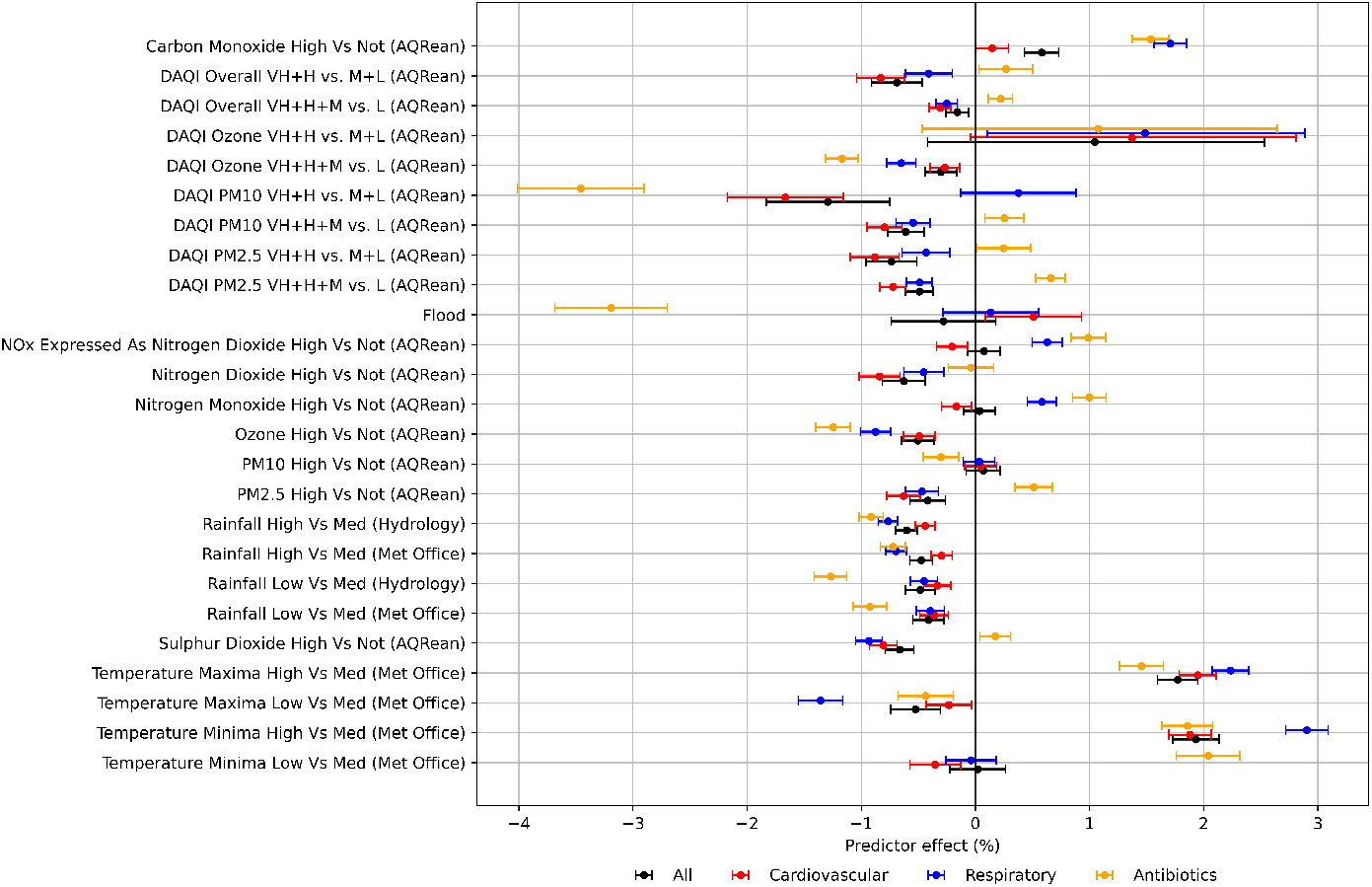
Coefficient estimates from mixed-effects model for change in prescription volume (as percentages) for flagged months. Error-bars show 95% confidence intervals. Sulphur dioxide daily air quality index (DAQI) flags have been excluded as a lack of variability led to large errors, reducing visibility of the effects for other flags. Results for nitrogen dioxide DAQI flags were also excluded as all periods were categorised as low (DAQI = 1–2). Nox = nitrous oxides. PM2.5 and PM10 refer to particulate matter < 2.5*µ*m and < 10*µ*m in diameter, respectively. VH, H, M, L refer to “very high”, “high” “moderate” and “low” DAQI categories, respectively.

High pollutant concentrations were generally associated with minimal changes in prescription volumes. Periods of elevated carbon monoxide concentrations showed the strongest associations, with small increases in cardiovascular medications and 1.4–1.9% increases in antibiotic and respiratory prescriptions. Comparisons across daily air quality index (DAQI) categories similarly indicated minimal associations overall. However, when comparing “very high” and “high” PM_10_ DAQI categories to “moderate” and “low”, a strong negative association was observed (−2.9 to −4%). This effect was not apparent when comparing “low” to all other categories, and 95% confidence intervals were wide due to the small number of months flagged in the “high” and “very high” categories relative to “low” and “moderate”. Ozone concentrations also showed some association with antibiotic prescriptions, with effects of −1 to −1.4% for both concentration-based and DAQI “low” versus all other category comparisons.

Overall, these results suggest that extreme environmental conditions do not lead to large, abrupt changes in monthly prescribing volumes, with most observed effects remaining modest in magnitude.

#### Relative importance of temperature, rainfall, and pollution

To assess how prescribing responds to incremental changes in environmental exposures, we next fitted mixed-effect models using standardised continuous predictors. Expressing effects per standard deviation increase enables direct comparison across variables measured on different scales and provides insight into the relative importance of temperature, precipitation and air-pollution measures.

Results for all continuous predictors (including DAQI values) are summarised in Figure 3, with effects expressed as the average percentage change in prescriptions associated with a standard deviation increase in each predictor. The strongest associations were observed for monthly temperature minima and maxima, which showed positive associations for most medication types. Increases in both minimum and maximum temperatures were associated with higher prescription volumes for respiratory (1.0–1.8%) and cardiovascular (0.6–0.9%) medications. Antibiotics, however, exhibited little association with minimum temperatures (−0.1 to 0.3%) and a more pronounced negative association with maximum temperatures (−1.1 to 1.5%). Monthly rainfall showed minimal impact on prescription volumes across all categories, with all effect estimates below 0.3%.

**Figure 3.**
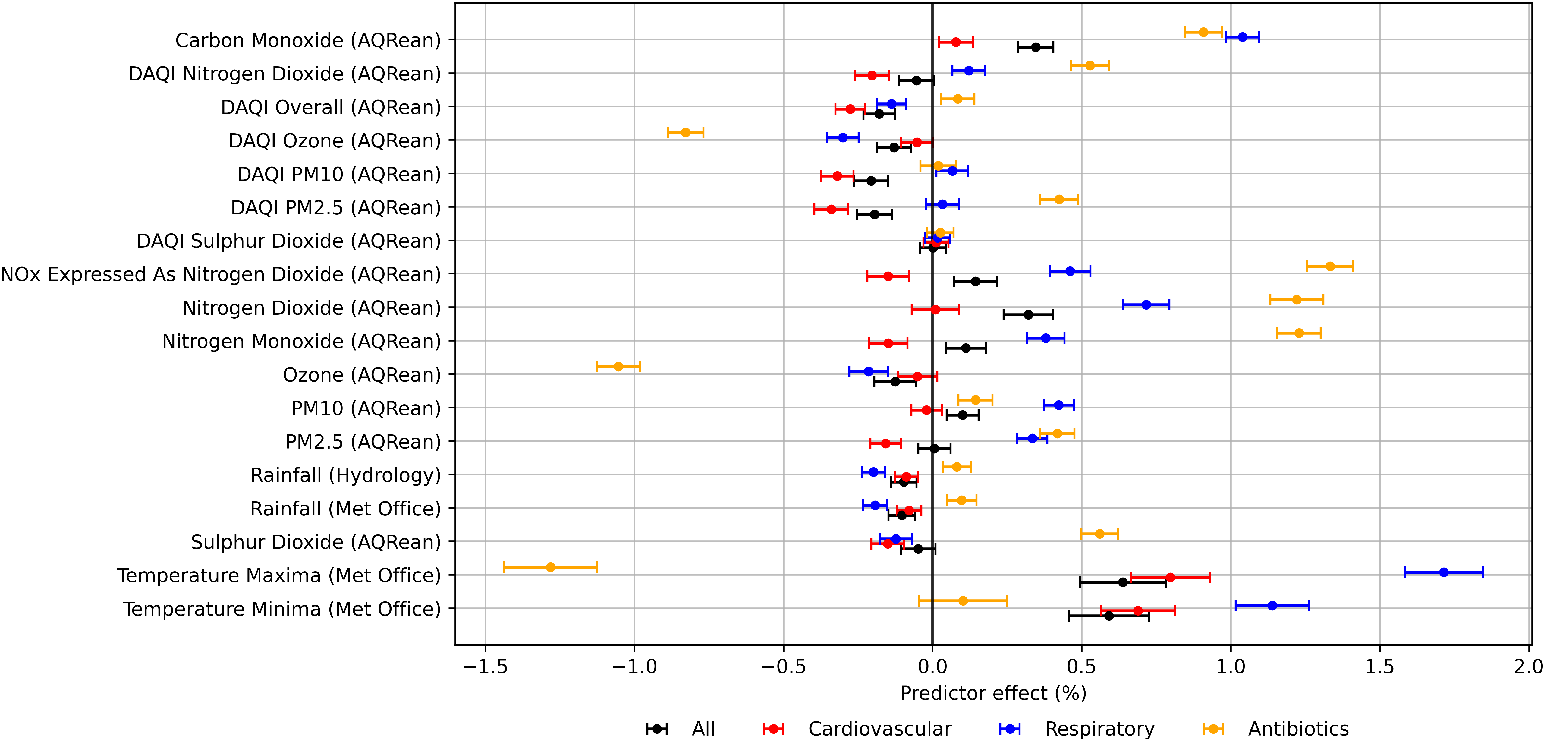
Coefficient estimates from mixed-effects models for change in prescription volumes (in percentages) relative to standard deviation increases in predictor values. Error-bars show 95% confidence intervals. NOx = nitrous oxides. PM2.5 and PM10 refer to particulate matter < 2.5*µ*m and < 10*µ*m in diameter, respectively.

While most air-quality predictors showed minimal associations (< 0.5%), carbon monoxide and ozone exhibited effects similar to those observed for anomalous periods. Increases in carbon monoxide concentrations were associated with a very small rise in cardiovascular prescriptions (< 0.2%), but antibiotics and respiratory medications increased by 0.8–1.2% per standard deviation increase in carbon monoxide. Ozone increases were generally associated with decreases in antibiotics prescriptions, with effects of approximately −0.8% for DAQI category increases and −1.1% for concentration increases. Nitrogen-based particulates also showed a stronger effect on antibiotics prescriptions (1.1–1.4%) compared to the flagged month comparisons.

These results indicate that temperature-related variables, rather than rainfall or most pollutants, show the most consistent associations with prescribing across medication groups when considering typical month-to-month variation.

#### Socioeconomic and geographic drivers of prescribing

Socioeconomic deprivation and geographic region are well-established determinants of prescribing behaviour and healthcare utilisation. Although not the primary focus of this study, examining these effects provides important context for interpreting environmental associations and helps quantify their relative magnitude.

Standard deprivation and regional effects were estimated in all models, but results are not shown due to the large number of models. Increases of one standard deviation in local area Index of Multiple Deprivation (IMD) percentiles (indicating lower deprivation) were weakly associated with respiratory prescriptions (0–6%) and more strongly associated with higher cardiovascular and antibiotic prescribing (5–15%).

Most region-level mean effects were close to zero, with 95% confidence intervals frequently spanning zero, suggesting little evidence of meaningful differences in prescribing between regions after accounting for all other covariates. However, a few regions exhibited more pronounced effects. London showed negative associations with cardiovascular (−26 to 39%), respiratory (−35 to 42%), and antibiotic (−25 to 32%) prescribing. In contrast, the North East exhibited 5–50% higher cardiovascular prescribing. Several other regions, including the East Midlands, South West, and Yorkshire and the Humber, also demonstrated higher prescribing of cardiovascular and respiratory medications of up to 40%, whereas the West Midlands and South East were up to 25% below the national average.

The substantially larger effects associated with deprivation and region underscore that environmental influences on prescribing operate within broader structural and socioeconomic contexts.

### 3.3 Multivariate analysis of environmental predictors

While individual mixed-effects models provide interpretable marginal associations, they cannot disentangle the conditional effects of correlated environmental exposures. We therefore fitted a Bayesian hierarchical multivariate model incorporating multiple predictors simultaneously. This approach allowed us to estimate adjusted associations while accounting for collinearity and propagating uncertainty, with partial pooling applied to practice-level effects to stabilise estimates for smaller practices.

Posterior estimates from the Bayesian multivariate models are presented in Figure 4. Temperature exhibited the strongest associations among all predictors, with higher maximum temperatures associated with increased respiratory prescription volumes (2.7–3.1%) and smaller positive effects for total prescriptions and cardiovascular medications. In contrast, higher maximum temperatures were associated with reductions in antibiotic prescribing (1.6– 2.1%). Minimum temperature showed consistently small negative associations across medication categories (− 0.1 to −1.3%), with the largest effect observed for antibiotics. The immediate effects of maximum temperature were consistent between lagged and non-lagged models across all prescription categories (results presented in Appendix A, Figure A1). At a two-month lag, however, associations with cardiovascular and respiratory prescriptions became negative, while the effect on antibiotic prescriptions attenuated towards zero. In contrast, minimum temperature exhibited positive associations with all prescription types, with effect estimates increasing in magnitude at longer lags. Rainfall demonstrated near-zero effects across all medication types, and the flood indicator likewise showed only weak positive associations (0.0–1.0%) with wide credible intervals, indicating minimal evidence of a meaningful relationship with prescription volumes. Rainfall effects appeared to become stronger at longer lags, but the effects were still small.

**Figure 4.**
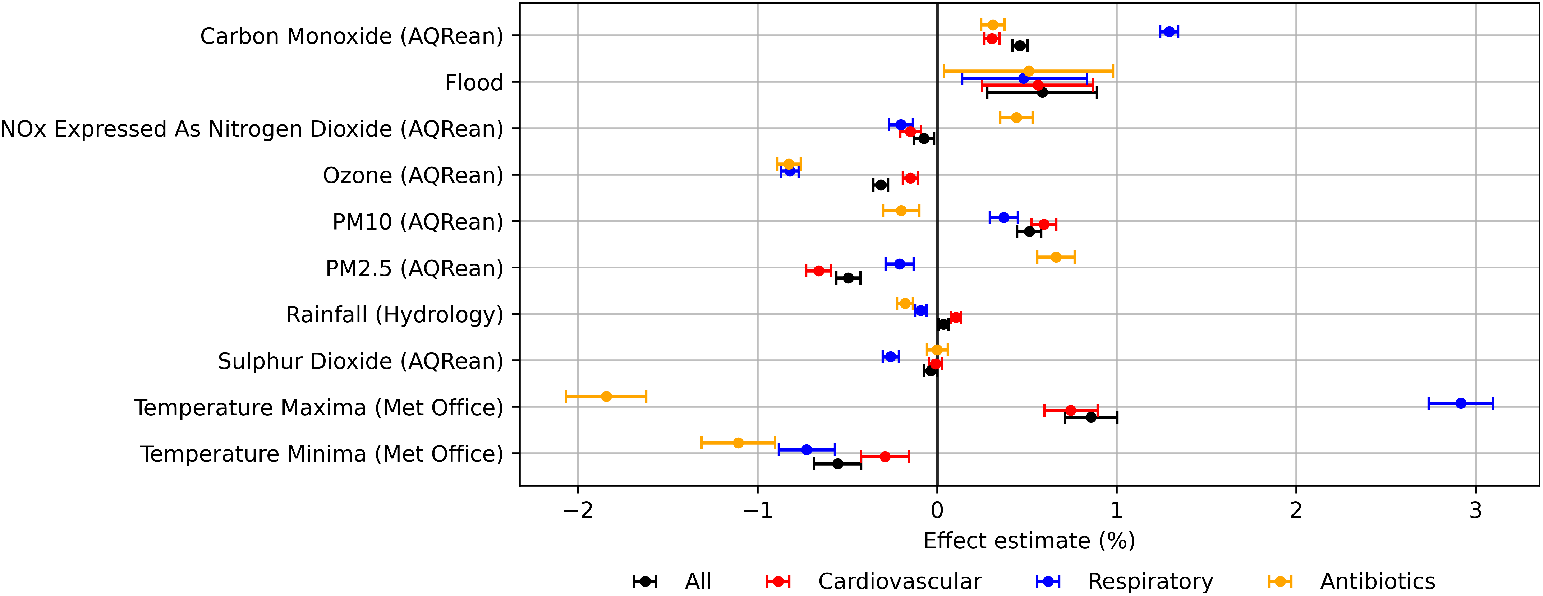
Posterior estimates from Bayesian multivariate model for percentage change in prescription volumes relative to standard deviation increase in predictors. Error-bars show 95% equal-tailed credible intervals. NOx = nitrous oxides. PM2.5 and PM10 refer to particulate matter < 2.5*µ*m and < 10*µ*m in diameter, respectively.

Pollutant concentrations were generally associated with small effect sizes, with both the magnitude and direction of associations varying across individual compounds. Carbon monoxide showed weak positive associations across most medication categories, with a stronger positive effect of 1.2–1.3% on respiratory prescriptions. Nitrogen-based pollutants exhibited minimal associations overall, although a modest positive relationship with antibiotic prescriptions was observed (0.4–0.5%). Ozone was associated with small reductions in prescription volumes, with negligible effects on cardiovascular medications but approximately 0.9% reductions in respiratory prescriptions and antibiotics. Sulphur dioxide demonstrated no substantive association with any prescription category.

Associations for particulate matter differed by particle size and prescription category. PM_2.5_ showed weak negative associations with cardiovascular and respiratory prescriptions (−0.1 to −0.7%) and a small positive association with antibiotic use (0.6–0.8%), whereas PM_10_ demonstrated the opposite pattern, with weak positive associations for cardiovascular and respiratory medications (0.3–0.6%) and a small negative association for antibiotics (−0.1 to −0.3%. The results from the lagged model did not support any substantial change in effects over time for pollutant and air quality measures.

Taken together, the multivariate results indicate that temperature remains the dominant environmental predictor of prescribing, while the independent contributions of rainfall, flooding and most pollutants are small once shared seasonal and meteorological structure is accounted for.

#### Regional and socioeconomic effects

The effects of the Index of Multiple Deprivation (IMD), expressed as the percentage change in prescription volumes associated with a one standard deviation increase in regionally centred IMD percentiles, are presented in Figure 5.

**Figure 5.**
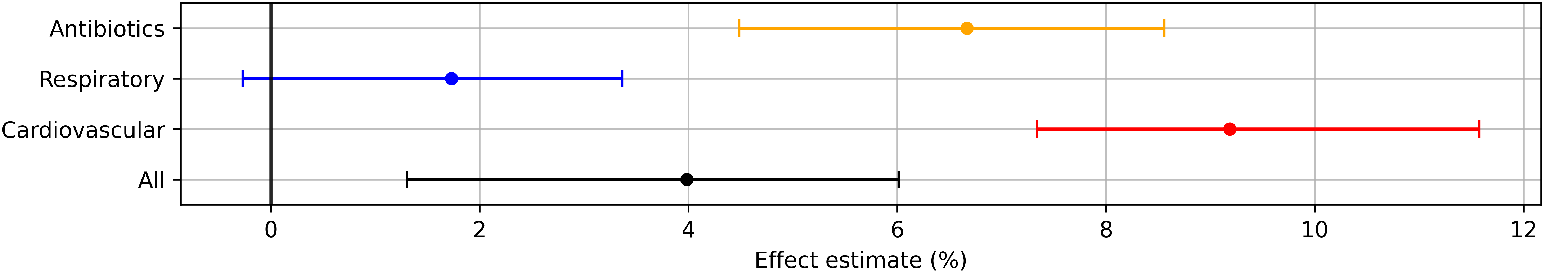
Percentage change in prescription volumes relative to global standard deviation increase in region-centred Index of Multiple Deprivation percentiles (decreasing deprivation within region).

Overall, prescriptions tended to increase in wealthier areas for antibiotics (4.5–8.5%) and cardiovascular (7.3–11.5%) medications, with a smaller increase observed for respiratory medications (−0.2–3.3%). However, the 95% credible intervals are wide, and model diagnostics indicated some difficulty in estimating the IMD effect. These results should therefore be interpreted cautiously.

Regional differences relative to the national mean are shown in Figure 6 and are consistent with the results of the mixed-effects models. Prescription volumes for all three medication types were 26–41% below the national mean in London. The West Midlands had 3–23% lower prescribing, while the South East showed up to 12% fewer respiratory and cardiovascular prescriptions but 8–18% higher antibiotic prescriptions. In contrast, prescribing tended to be above the national mean in other regions. The East of England and South West had higher prescribing across all medication types (up to 30%), cardiovascular prescribing was elevated in the North East (9–35%), Yorkshire and The Humber (3–14%), and East Midlands (9–21%), and respiratory prescribing exceeded the national mean in the North West (5–15%), Yorkshire and The Humber (6–18%), and East Midlands (12–20%).

**Figure 6.**
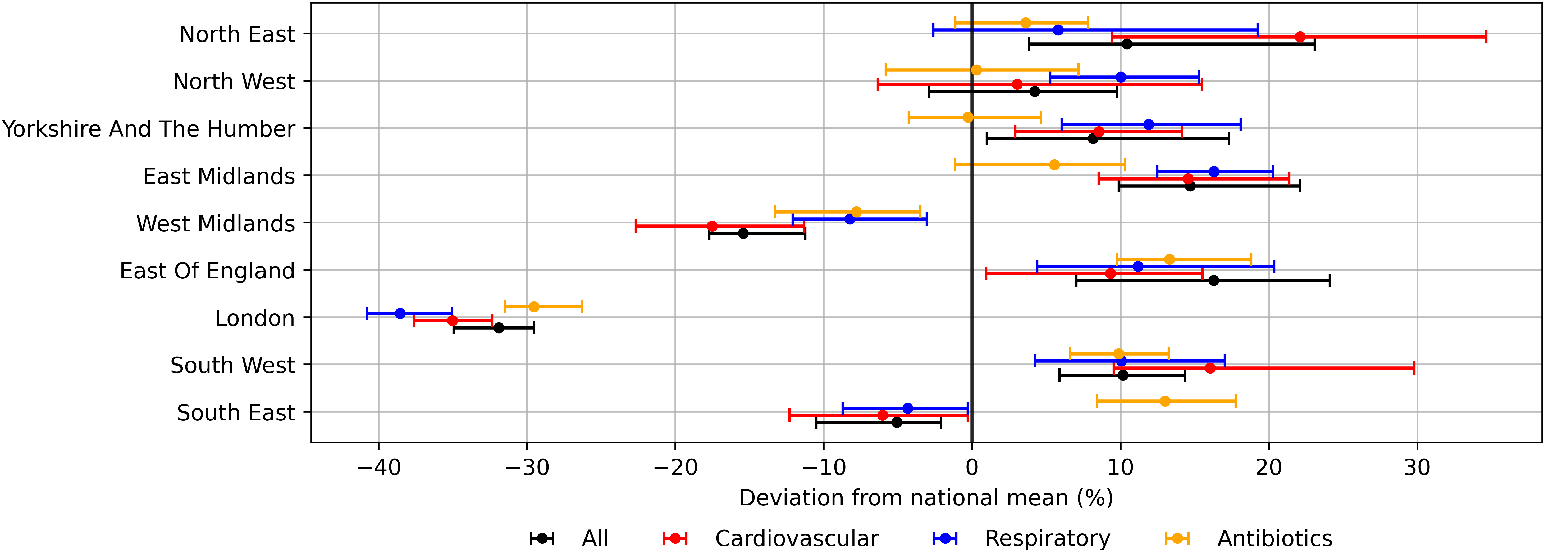
Regional variation in prescription uptake in comparison to the national mean across England. Both panels include error-bars to represent 95% equal-tailed credible intervals and follow the same colour-coding for prescription types as shown in the legend at the bottom of the figure.

## 4 Discussion

This study examined associations between climate variables, air pollution and GP prescription uptake in the UK (England) using large-scale, longitudinal prescribing data linked to multiple environmental datasets. Across modelling approaches, temperature emerged as the most consistent environmental predictor of prescribing variation, while rainfall, flooding and most air pollutants showed weak or inconsistent associations once shared seasonal and meteorological structure was accounted for. Notably, environmental effect sizes were modest compared with socioeconomic and regional determinants of prescribing, indicating that environmental variability operates within a broader structural context shaping healthcare utilisation.

Temperature-related associations persisted across modelling approaches, but their interpretation depended on accounting for correlated exposures and seasonality. Divergent effects of minimum and maximum temperature in multivariate models suggest that univariate associations may partly reflect shared seasonal structure rather than distinct mechanisms. These findings are consistent with existing evidence that temperature influences cardiovascular and respiratory health, while also indicating that prescribing responses are nuanced and dependent on seasonal context rather than simple linear exposure–response relationships [8]. The consistent negative association between temperature and antibiotic prescribing, despite the positive association between temperature and infection risk reported elsewhere [5], likely reflects reduced winter infection burden and seasonal healthcare-seeking behaviour, rather than direct temperature effects alone.

In contrast, precipitation and flooding showed little evidence of meaningful association with prescribing volumes at monthly timescales. Apparent flooding effects observed in univariate models were not supported after adjustment for correlated environmental predictors, suggesting that these associations may reflect confounding rather than direct impacts on prescribing behaviour. The rarity of flood events within the study period further limited power to detect small effects, and short-term disruptions may not be captured in monthly prescribing data.

Associations between air pollution and prescribing were generally weak and sensitive to model specification. Carbon monoxide was the most consistent pollutant predictor, although effect sizes were small, while nitrogen-based pollutants, particulate matter and ozone showed weak or inconsistent associations once correlations were accounted for. Although carbon monoxide is projected to decline under most future scenarios, increases remain possible under high-emissions pathways, and climate-driven wildfires may lead to episodic local elevations in particulate matter and carbon monoxide [21, 22, 23]. Taken together, these findings suggest that while air pollution contributes to morbidity, its influence on monthly prescribing demand is likely to be modest on average, with potential for short-term localised impacts under extreme conditions.

Socioeconomic deprivation and geographic region were strongly associated with prescribing across all models, with substantially larger effect sizes than those observed for environmental predictors. This underscores that environmental influences on prescribing are embedded within persistent structural and socioeconomic determinants of healthcare utilisation, and that climate-related effects should be interpreted in this broader context.

The principal strength of this study lies in its integration of nationwide prescribing data from over 15,000 GP practices with multiple high-resolution environmental datasets, combined with modelling approaches that explicitly address correlated exposures. Limitations include sparse flood exposure, potential exposure misclassification from station-based and gridded data, monthly aggregation of prescribing outcomes that may obscure short-term effects, and the possibility of residual confounding from unmeasured behavioural or healthcare system factors.

Overall, these findings suggest that GP prescribing volumes are relatively stable at monthly timescales in response to environmental variability. While temperature-related changes may have cumulative implications for healthcare demand under long-term climate change, rainfall, flooding and most air pollutants appear to play a limited role once broader temporal structure is accounted for. This supports a focus on long-term adaptation and surveillance, rather than short-term demand shocks, in planning climate-resilient healthcare systems.

From a public health perspective, these findings suggest that monthly prescribing demand in primary care is relatively stable to short-term environmental variability. Rather than anticipating acute surges in prescribing associated with individual weather or pollution events, climate-resilient healthcare planning may benefit more from long-term surveillance of temperature-related trends alongside persistent socioeconomic and regional drivers. Integrating routine climate indicators into prescribing surveillance systems could support early identification of gradual shifts in demand under future climate change.

## 5 Conclusions

Our findings suggest that temperature is the most consistent climatic determinant of prescribing behaviour, with higher maximum monthly temperatures associated with increased cardiovascular and respiratory prescribing and warmer temperature minima linked to modest reductions. Rainfall and flooding showed little evidence of meaningful effects once seasonal and meteorological confounding were accounted for. Most pollutants displayed weak or inconsistent associations, although carbon monoxide emerged as a notable predictor of increased prescribing across multiple medication groups, predominantly for respiratory medications. These results suggest that, at monthly timescales, prescribing demand in primary care is relatively robust to environmental variability. For public health planning, this supports an emphasis on long-term surveillance and adaptation to gradual temperature-related changes rather than short-term demand shocks linked to individual environmental events.

Future work could utilise higher-resolution prescribing and environmental data to investigate short-term acute effects, and to examine how environmental exposures interact with deprivation and other socioeconomic factors to influence prescription volumes. As climate and air quality evolve in the coming decades, further research and continued monitoring will be important for anticipating their implications for healthcare demand.

## Data Availability

All data produced in the present study are available upon reasonable request to the authors.

## 6 List of abbreviations

API: Application Programming Interface
AQREAN: Air Quality REANalysis
BNF: British National Formulary
DAQI: Daily Air Quality Index
DEFRA: Department for Environment, Food & Rural Affairs
GP: General Practitioner
IMD: Index of Multiple Deprivation
LAD: Local Authority District
MAD: Median Absolute Deviation
NHS: National Health Service
PM: Particulate Matter
UK: United Kingdom

## 7 Declarations

### Ethics approval and consent to participate

Not applicable: all data were collected from open-source platforms and completely anonymised.

### Consent for publication

Not applicable: no data relating directly to individuals was used in this study.

### Availability of data and materials

Prescription data is available from Open Prescribing and NHS Digital [20]. Weather related data is available from Met Office [13] and Department for Environment Food & Rural Affairs Hydrology API [14]. Flood data is available from UK Environment Agency [15]. Air quality data is available from UK Air Quality Reanalysis [16]. Regional and deprivation data are available from Office for National Statistics [17] and an array of government departments [18]. Results generated in this study are available on reasonable request.

### Authors’ contributions

JT collected and pre-processed datasets, and ran the analyses. JT and CY interpreted the results, and wrote and edited the manuscript. CY provided funding and supervision. All authors read and approved the final manuscript.

## Acknowledgements

Not applicable.

## Appendices

### A Lagged environmental effects

**Figure A1.**
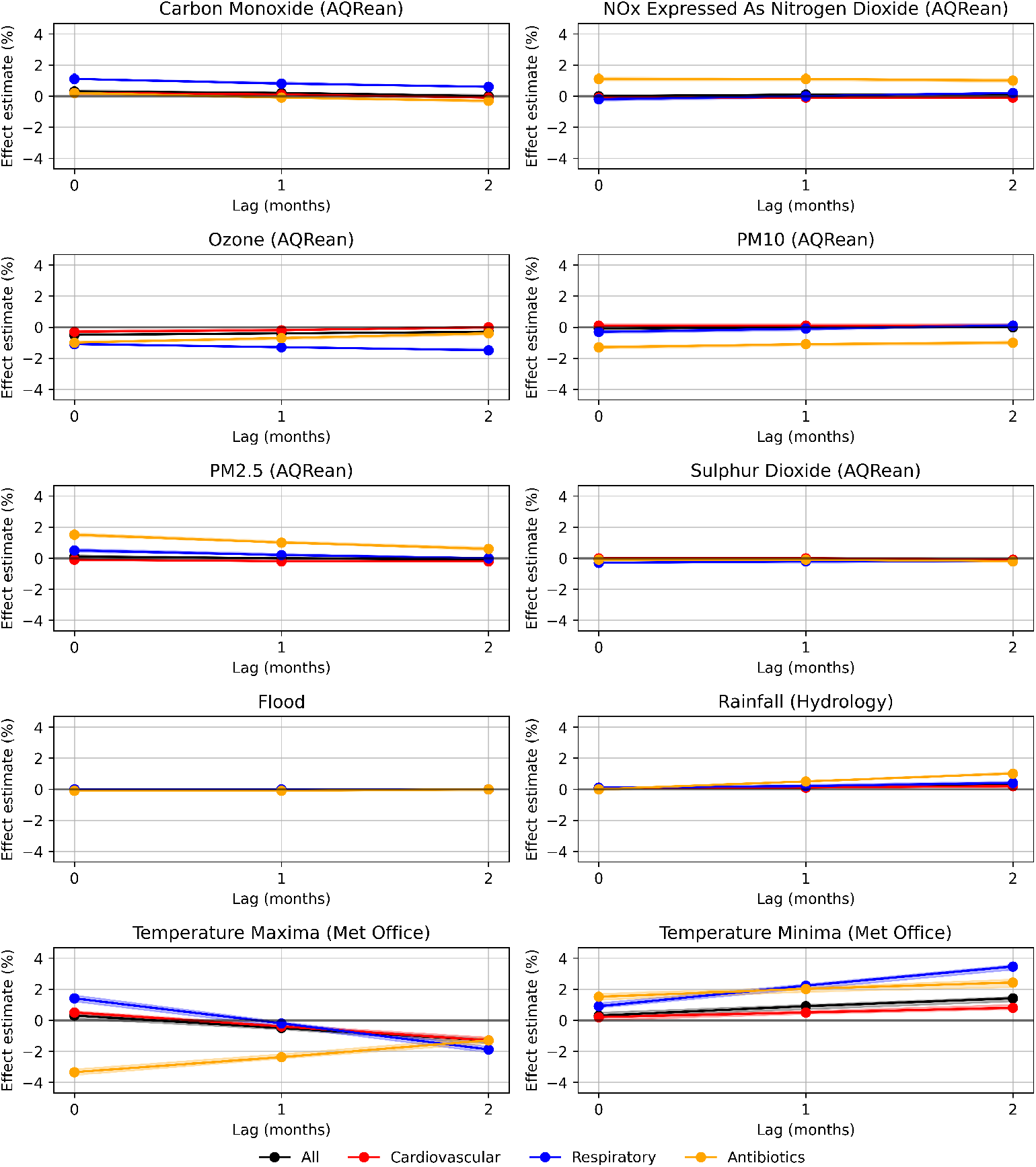
Posterior estimates from Bayesian multivariate model for percentage change in prescription volumes relative to standard deviation increase in predictors with up to two months lag. Shading around each line shows 95% equal-tailed credible intervals. Nox = nitrous oxides. PM2.5 and PM10 refer to particulate matter < 2.5*µ*m and < 10*µ*m in diameter, respectively.

